# Multimodal Diverse Granularity Fusion Network based on US and CT Images for Lymph Node Metastasis Prediction of Thyroid Carcinoma

**DOI:** 10.1101/2023.12.25.23300117

**Authors:** Guojun Li, Jincao Yao, Chanjuan Peng, Yinjie Hu, Shanshan Zhao, Xuhan Feng, Jianfeng Yang, Dong Xu, Xiaolin Li, Chulin Sha, Min He

## Abstract

Accurately predicting the risk of cervical lymph node metastasis (LNM) is crucial for surgical decision-making in thyroid cancer patients, and the difficulty in it often leads to over-treatment. Ultrasound (US) and computed tomography (CT) are two primary non-invasive methods applied in clinical practice, but both contain limitations and provide unsatisfactory results. To address this, we developed a robust and explainable multimodal deep-learning model by integrating the above two examinations. Using 3522 US and 7649 CT images from 1138 patients with biopsy-confirmed LNM status, we showed that multimodal methods outperformed unimodal counterparts at both central and lateral cervical sites. By incorporating a diverse granularity fusion module, we further enhanced the area under the curve (AUC) to 0.875 and 0.859 at central and lateral cervical sites respectively. This performance was also validated in an external cohort. Additionally, we quantified the modality-specific contributions for each nodule and systematically evaluated the applicability across various clinical characteristics, aiding in identifying individuals who can benefit most from the multimodal method.

---

The global incidence of thyroid cancer has surged over the past 30 years[1], reaching over 586,000 new cases in 2020[2]. Despite its generally indolent nature, thyroid cancer leads to cervical lymph node metastasis (LNM) in up to 50% of patients[3]. Cancer cells typically initially metastasize to the central lymph nodes and subsequently spread to the lateral cervical site, increasing the risk of recurrence and poor prognosis[4]. Consequently, LNM status significantly influences the surgical approach for thyroid cancer patients. Therapeutic lymph node dissection (LND) of central and lateral cervical compartments is normally recommended for individuals with central and/or lateral cervical LNM[5]. While for patients without LNM, although central LND remains controversial, prophylactic lateral cervical LND is not advised[5, 6]. However, the current non-invasive diagnostic accuracy of LNM is insufficient to guide surgical decisions. For the central site, the primary imaging methods, including Ultrasound (US) and computed tomography (CT), provide average sensitivities of only 0.28 and 0.39[7], respectively. This leads to a prevalent tendency for overtreatment to prevent missed LNM detection and results in potential complications such as recurrent laryngeal nerve injury and hypoparathyroidism. Therefore, there is a pressing need to improve the accuracy of LNM risk assessment to assist surgical management.

In recent years, the introduction of artificial intelligence methods has significantly improved the performance of LNM prediction. Several studies utilizing US images have employed various machine learning methods, such as gradient boosting, random forests, neural networks, etc., achieving AUCs in the range of 0.700 to 0.772[8, 9, 10] for predicting central site LNM. Other studies focusing on extracting high-dimensional radiomic features or employing deep learning methods to predict LNM status have achieved AUCs spanning from 0.78 to 0.90[11, 12, 13] for the central site and 0.62[14] for the lateral cervical site. Similarly, in the case of CT images, methods based on radiomic features extracted from thyroid nodules have demonstrated predictive capabilities for central site LNM at AUCs ranging from 0.710 to 0.770[15, 16].

However, it’s crucial to acknowledge that both US and CT modalities have limitations owing to their examination techniques. Though US images provide high-resolution visuals of thyroid nodules’ interior and boundary characteristics, their limited field of view poses challenges in assessing the spatial relationships between thyroid nodules and surrounding tissues. Conversely, CT images offer essential relative position information about the thyroid, lymph nodes, and surrounding tissues, albeit at a lower resolution compared to US images. Relying solely on unimodal methods restricts the predictive capabilities of the model. Considering that US and CT provide complementary information and are widely utilized in thyroid cancer diagnosis, there’s a great potential to improve the performance by integrating US and CT images through a multimodal approach for predicting LNM status. For instance, Zhao et al. developed a multivariate logistic regression multimodal model for predicting central LNM status by incorporating clinical factors, US-derived diagnostic features, and CT measurements, achieving an AUC of 0.827[17]. Nevertheless, the study did not directly compare the multimodal method’s performance with unimodal methods, besides, it utilized a simplified model that overlooked the interaction between the two modalities, leaving the full potential of multimodal fusion approach unexplored.

Leveraging deep learning methods for integrating multimodal medical data has emerged as a prominent approach to enhance our understanding of complex diseases[18, 19, 20], with promises in tailoring personalized diagnosis, prognosis, treatment, and care[21, 22, 23, 24]. The central premise of multi-modal data integration is that diverse data sources complement each other, augmenting information beyond any individual modality. However, significant challenges persist, such as data scarcity, sparsity, and inter-modality complexity, limiting the full exploitation of data integration benefits. Recent advancements in deep learning methods within this domain primarily focus on representation learning and fusion techniques[25, 26], which include extracting meaningful representations with unlabeled data[27, 28] and employing attention-based approaches to allow more sophisticated fusion of cross-modality representations[29, 30, 31, 32]. While these strategies exhibit improvements in model performance, their use in the biomedical field still requires broader testing across diverse scenarios and adaptation to specific tasks through dedicated study designs[33].

In this study, we aim to improve the predictive accuracy of LNM status for thyroid cancer patients by developing a multimodal method incorporating US and CT images. We curated a paired multi-modal dataset consisting of 3522 US and 7649 CT images from 1138 patients with biopsy-confirmed LNM status at both central and lateral cervical sites (Fig. 1). To comprehensively integrate the consistent and distinct information of both modalities, we first employed a multi-task network scheme to enhance modal-specific feature learning (Fig. 2), which achieved superior performance compared to currently commonly used methods on unimodal models. Next, we demonstrated that, even with a basic feature fusion strategy, multimodal models consistently outperform their unimodal counterparts at both sites. Furthermore, we designed a diverse granularity fusion module, which learns the attention at three granular levels from fine to coarse: dimension level, modality level, and nodule level (Fig. 2). With the incorporation of this module, our multimodal model achieved AUCs of 0.875 and 0.859 at the central and lateral cervical sites respectively. Compared to unimodal methods of US and CT, the multimodal AUC improved by 5.5% and 10.1% respectively, at the central compartment, and by 7.4% and 8.1% respectively, at the lateral cervical site. When evaluated on an external validation set, our proposed model demonstrated an AUC of 0.903 at the central site, which robustly confirmed the generalizability of the multimodal model. In addition, we comprehensively evaluate the applicability of each modality on nodules with various characteristics to identify patients who can best benefit from the multimodal method (Fig. 1), which could significantly improve the clinical utility of multimodal models. In summary, we presented a promising approach to mitigate the issue of overtreatment in thyroid cancer. Our multimodal AI system exhibits strong performance, high generalizability, and substantial clinical utility, offering significant potential for enhancing the diagnosis and treatment of thyroid cancer.

**Figure 1:**
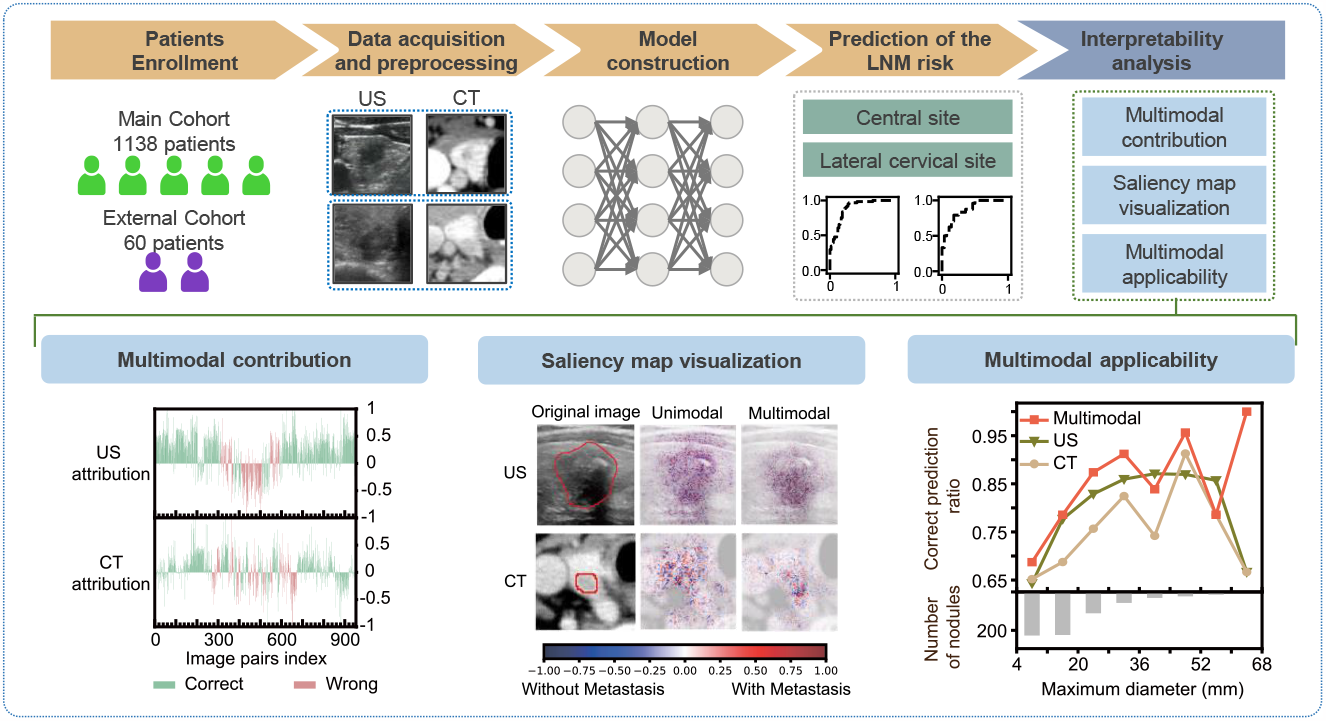
Overall AI system for LNM risk prediction. The main cohort was employed for AI system development and evaluation, while the external cohort assessed the system’s generalizability. After preprocessing, paired US and CT images are input into DGFNet, our deep learning model, to predict LNM status in central and lateral cervical regions. Post-AI system development, we conducted an extensive interpretability analysis comprising multimodal contribution assessment, saliency map visualization, and multimodal applicability evaluation.

**Figure 2:**
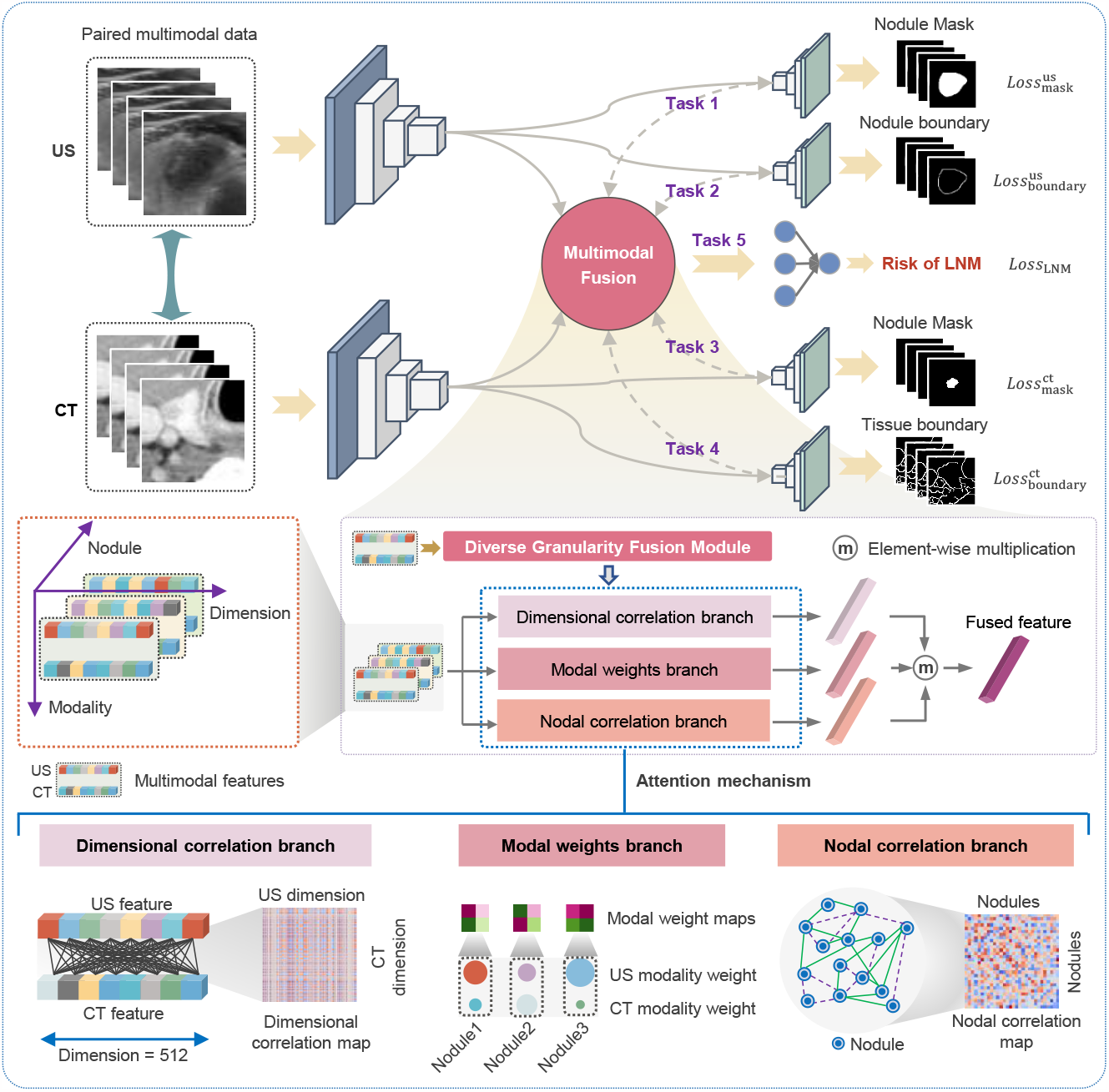
DGFNet architecture. DGFNet consists of three branches: the US branch, CT branch, and multimodal branch. Each US and CT branch incorporates an encoder and two decoders. DGFNet concurrently performs five tasks: nodal mask and boundary segmentation in US images (guiding the model to focus on internal and marginal nodule features), boundary segmentation of nodules and surrounding tissues in CT images (guiding the model to focus on nodule and surrounding tissue features in CT images), and the final LNM prediction. The fusion of multimodal features in the latent space occurs within the diverse granularity fusion module, and the final results are generated by subsequent fully connected layers. The diverse granularity fusion module includes the dimensional correlation branch, modal weights branch, and nodal correlation branch, amalgamating characteristics from both modalities to provide a diverse granularity information integration. A detailed explanation of this module is available in the Methods section.

## Results

### Patient Cohort

This study incorporated two datasets: a main cohort and an external cohort. The main cohort comprised patients who underwent thyroid examinations at Zhejiang Cancer Hospital from August 2018 to February 2021. To reflect the real clinical diagnostic conditions, only necessary data quality control was performed, with specific details outlined in the supplementary material. After quality control, the main cohort consists of 1138 patients with a total of 1285 thyroid nodules. The external cohort, obtained from Shaoxing People’s Hospital in Zhejiang Province, also underwent the same quality control process, comprising 60 patients with 60 thyroid nodules. Both cohorts included samples with matched US and CT data, featuring multiple images of thyroid nodules, along with their corresponding LNM status at the central and lateral cervical sites.

The models were evaluated under an eight-fold cross-validation setting, and various metrics were employed to assess their performance. These metrics encompassed accuracy (ACC), the area under the ROC curve (AUC), sensitivity (SENS), specificity (SPEC), precision (PREC), and the F1 score. We reported the mean metrics calculated from the eight-fold cross-validation process for a comprehensive evaluation. It is worth mentioning that samples from different folds were divided based on the individual thyroid nodules, and nodules from the same patient were consistently present within the same fold. In addition, an undersampling strategy was applied in this study to maintain a balance between positive and negative categories.

### Enhance modal-specific feature learning by employing multi-task models of each modality

We start from enhancing modal-specific feature extraction to make the best use of each modality and evaluate the feature capability in predicting LNM status of each modality. US provides clear visualization of thyroid nodule attributes such as boundary, shape, and internal structure (composition, calcification, echo characteristics). Meanwhile, CT images encompass both thyroid nodules and the surrounding anatomical context, offering insights into their relationships. Therefore, we employ a multi-task learning approach for each modality (as illustrated in Fig. 2). Specifically, besides the LNM prediction task, we introduce two auxiliary tasks for US: a nodule mask segmentation task to guide the model to focus on the internal structural features, and a nodule boundary segmentation task to emphasize the boundary and shape of nodules. Likewise, for CT, we introduce a nodule mask task and a tissue boundary segmentation task to guide the model to distinguish nodules and surrounding tissue regions, respectively. We chose ResNet[34] as the backbone to build multi-task models for each modality, due to its simple structure, high popularity, and excellent performance (Methods). For each unimodal network, we trained it to complete the auxiliary segmentation tasks in the first 100 epochs and added the additional LNM prediction task in the following 200 epochs. We compared our multi-task models for each modality with ResNet models directly predicting LNM, and for the US unimodal model, we also re-implemented the network developed by Yu et al[12]. It shows that our multi-task models for both modalities consistently outperform their counterparts at both central and cervical lateral sites, with an obvious improvement of ACC and AUC (Table 1).

**Table 1:** Performance of LNM status prediction using unimodal networks.

| Modalities | LNM location | Models | ACC | AUC | SENS | SPEC | PREC | F1 score |
| --- | --- | --- | --- | --- | --- | --- | --- | --- |
| US | Central site | Yu et al [12] | 0.739 | 0.792 | 0.787 | 0.689 | 0.719 | 0.750 |
|  |  | ResNet | 0.760 | 0.812 | 0.823 | 0.699 | 0.736 | 0.774 |
|  |  | Proposed | **0.782** | **0.820** | **0.836** | **0.728** | **0.754** | **0.792** |
|  | Lateral cervical site | Yu et al [12] | 0.729 | 0.754 | 0.729 | **0.731** | 0.736 | 0.728 |
|  |  | ResNet | 0.747 | 0.725 | 0.826 | 0.665 | 0.713 | 0.763 |
|  |  | Proposed | **0.767** | **0.785** | **0.843** | 0.693 | **0.737** | **0.784** |
| CT | Central site | ResNet | 0.726 | 0.759 | **0.743** | 0.707 | 0.720 | 0.730 |
|  |  | Proposed | **0.745** | **0.774** | 0.737 | **0.755** | **0.751** | **0.743** |
|  | Lateral cervical site | ResNet | 0.746 | 0.758 | **0.812** | 0.679 | 0.718 | **0.760** |
|  |  | Proposed | **0.764** | **0.778** | 0.725 | **0.797** | **0.796** | 0.751 |

When comparing the unimodal performance of US and CT, we have some interesting observations. First, at the central site, the US models generally outperform CT models, whereas there is no consistent winner between US and CT models at the lateral cervical site. Moreover, at both the central and lateral cervical sites, US models consistently exhibit higher sensitivity, meanwhile, CT models consistently demonstrate higher specificity. These results suggest that US is more sensitive but less specific, while CT is the opposite, highlighting the complementary information provided by these two modalities.

### Basic multimodal fusion methods outperform either unimodal model

Based on the multi-task unimodal models, we further evaluate the efficacy of integrating US and CT for predicting the risk of LNM. We first examine the multimodal performance using three basic fusion methods: concatenation, element-wise sum, and element-wise multiplication, to fuse the unimodal features extracted from US and CT and re-train the multimodal network in an end-to-end manner. The results clearly show that even with basic fusion methods, multimodal models significantly improve performance.

For the central site prediction, the average multi-modal AUC improved by 2.8% and 7.4% compared to US and CT unimodal respectively. Likewise, for the lateral cervical site prediction, the average multimodal AUC outperforms the US and CT unimodal models by 5.3% and 6.0% respectively (Table 2). These results affirm the hypothesis that US and CT modalities comprise complementary information, and their integration can improve the performance of LNM status prediction.

**Table 2:** Performance comparison of unimodal and multimodal approaches using basic fusion methods.

| LNM location | Modality | Multimodal fusion operation | ACC | AUC | SENS | SPEC | PREC | F1 score |
| --- | --- | --- | --- | --- | --- | --- | --- | --- |
| Central site | US |  | 0.782 | 0.820 | 0.836 | 0.728 | 0.754 | 0.792 |
|  | CT |  | 0.745 | 0.774 | 0.737 | 0.755 | 0.751 | 0.743 |
|  | Multimodal | Concat | 0.810 | **0.853** | **0.862** | 0.759 | 0.782 | 0.819 |
|  |  | Sum | 0.806 | 0.850 | 0.855 | 0.757 | 0.779 | 0.814 |
|  |  | Product | **0.812** | 0.840 | 0.861 | **0.763** | **0.790** | **0.820** |
|  |  | Average results | 0.809 | 0.848 | 0.859 | 0.760 | 0.784 | 0.818 |
| Lateral cervical site | US |  | 0.767 | 0.785 | 0.843 | 0.693 | 0.737 | 0.784 |
|  | CT |  | 0.764 | 0.778 | 0.725 | **0.797** | **0.796** | 0.751 |
|  | Multimodal | Concat | 0.804 | 0.825 | 0.860 | 0.744 | 0.779 | 0.815 |
|  |  | Sum | 0.808 | **0.854** | **0.878** | 0.742 | 0.771 | 0.819 |
|  |  | Product | **0.811** | 0.835 | 0.867 | 0.755 | 0.784 | **0.821** |
|  |  | Average results | 0.808 | 0.838 | 0.868 | 0.747 | 0.778 | 0.818 |

### Further improve multimodal performance by incorporating a diverse granularity feature fusion module

Multimodal fusion using basic methods can combine US and CT information and improve LNM prediction but is not able to fully consider the interaction between these two modalities. Recent progress based on the attention mechanism has shown superiority in multimodal fusion. In our study, we adopted the attention mechanism simultaneously on three granular levels to fully incorporate the information useful for LNM prediction, which are feature dimensions level (minimum granularity), modalities level (medium granularity), and nodules level (maximum granularity). In specific, these include dynamically adjusting the attention weights of different feature dimensions to balance the common and specific features of the two modalities, adapting the modality-specific attention to learn the respective advantages for different nodules, plus flexibly aggerate the features of other nodules based on nodule-level attention to refine the prediction, considering nodules with the same LNM status should exhibit greater feature similarity. We refer to our modality fusion methods as the ‘diverse granularity fusion’ network (DGFNet, as illustrated in Fig. 2, detail see Methods). Equipped with the DGF module, our model demonstrates exceptional predictive capabilities with AUCs of 0.875 and 0.859 at the central and lateral cervical sites respectively (Table 3), indicating its remarkable performance in predicting the risk of LNM. Particularly, the multi-modal AUC exhibited a significant improvement of 5.5% and 10.1% compared to the US and CT uni-modal models at the central site, respectively. And a substantial enhancement of 7.4% and 8.1% respectively at the lateral cervical site. These results further underscore the efficacy of integrating US and CT in predicting LNM in thyroid cancer. Furthermore, in comparison to the basic fusion methods, our DGFNet model achieves superior performance in nearly all metrics, providing comprehensive evidence for the effectiveness of fusing multimodal features at different granularities.

**Table 3:** Performance comparison of multimodal methods using different fusion techniques.

| LNM location | Multimodal fusion methods | ACC | AUC | SENS | SPEC | PREC | F1 score |
| --- | --- | --- | --- | --- | --- | --- | --- |
| Central site | Basic fusion | 0.809 | 0.848 | **0.859** | 0.760 | 0.784 | 0.818 |
|  | Diverse granularity fusion | **0.826** | **0.875** | 0.848 | **0.803** | **0.813** | **0.830** |
| Lateral cervical site | Basic fusion | 0.808 | 0.838 | **0.868** | 0.747 | 0.778 | 0.818 |
|  | Diverse granularity fusion | **0.838** | **0.859** | 0.862 | **0.814** | **0.835** | **0.842** |

### DGFNet demonstrates exceptional generalization abilities

Recognizing the importance of the generalizability of multimodal networks in clinical applications, we evaluated the efficacy of our DGFNet model using an external test dataset. The primary cohorts were partitioned into training and validation sets, and the model with the highest accuracy on the validation set was selected to predict the LNM status of patients on the external cohort. Owing to constraints related to data availability, our external evaluation is only performed at the central site, the results are presented in Table 4. Overall, our DGFNet model performed well on the external dataset, with an accuracy of 0.817 and an AUC of 0.903, showing similar performance compared to the internal accuracy of 0.844 and an AUC of 0.898. This consistency underscores the strong robustness and external generalizability of our model.

**Table 4:** Performance of LNM status prediction on internal and external validation set

| Dataset | ACC | AUC | SENS | SPEC | PREC | F1 score |
| --- | --- | --- | --- | --- | --- | --- |
| Internal validation set | 0.844 | 0.898 | 0.854 | 0.833 | 0.837 | 0.845 |
| External validation set | 0.817 | 0.903 | 0.909 | 0.763 | 0.690 | 0.784 |

### DGFNet dynamically adjusts the contribution of US and CT in predicting the LNM status prediction at different sites

We next seek to delineate the contribution of each modality in the DGFNet model on every nodule. We analyze by quantifying the contributions of US and CT within the DGFNet model using the integrated gradients[35] and comparing them to their unimodal counterparts. The results are presented in Fig. 3, where a larger feature attribution value corresponds to a greater contribution to the correct prediction in DGFNet model, and the red or green denotes correct or wrong prediction respectively.

**Figure 3:**
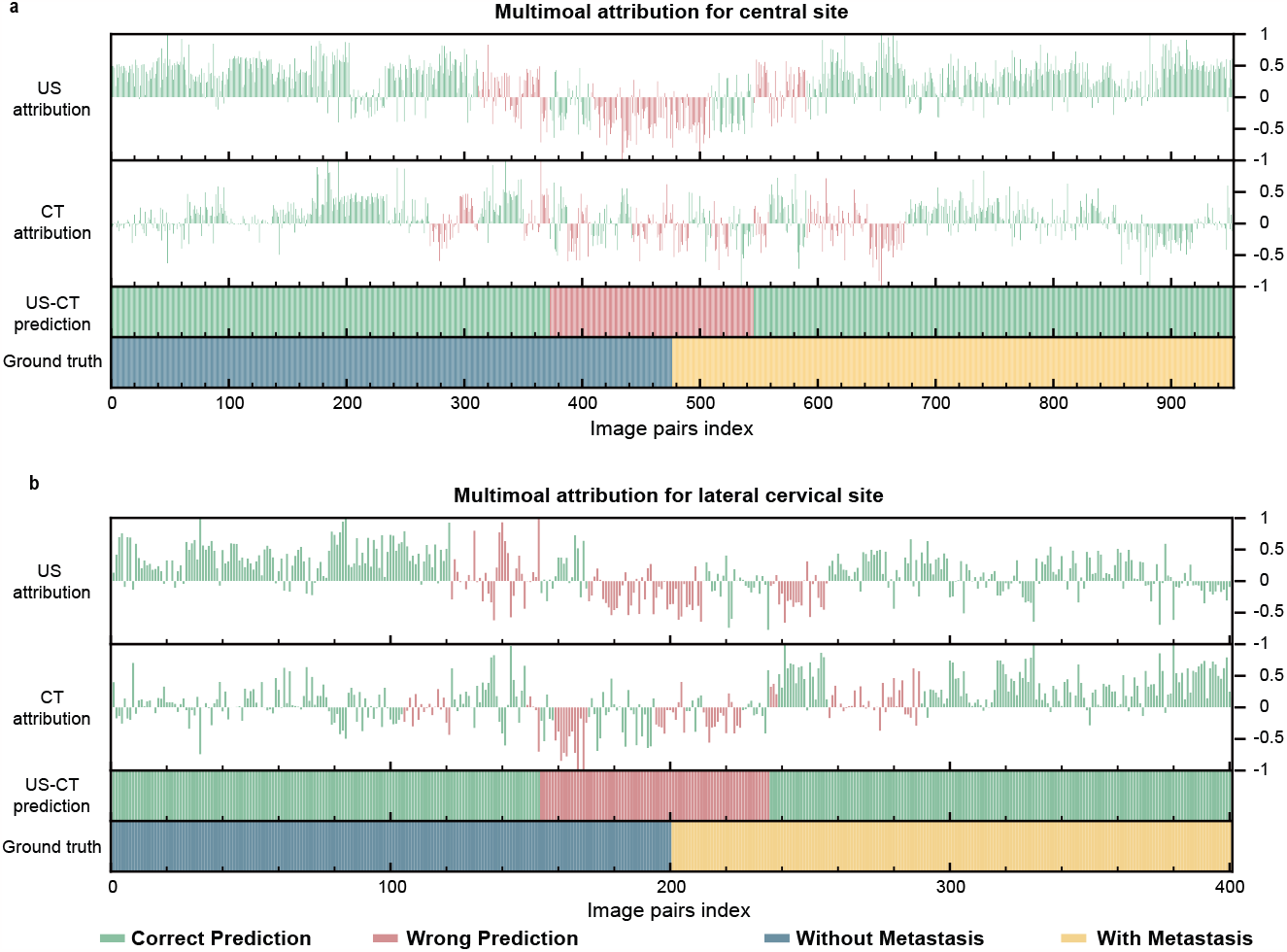
Attribution analysis of US and CT in predicting LNM status at central (a) and lateral cervical (b) sites. Each subfigure comprises four panels, with the shared horizontal axes indicating nodule indices. The central site includes 954 nodules, while the lateral cervical site includes 402 nodules. The values of the top two panels display attributions from US and CT images in the multimodal prediction, respectively. Column colors denote unimodal predictions, where green signifies accurate predictions and red indicates inaccuracies. The third panel illustrates the multimodal prediction results. Panel 4 represents the ground truth.

The result shows that, at both the central site (Fig. 3a) and lateral cervical site (Fig. 3b), there is a notable number of cases where the DGFNet can change the unimodality to make positive contributions even when it fails to give correct prediction in unimodal models (attribution greater than 0 but red color) and lead to a correct prediction in this multimodal approach. In addition, when looking at the central and lateral cervical sites separately, we find that, across all samples, 64.9% of nodules exhibit higher US attribution over CT attribution at the central site, while 55.2% of the nodules show higher US attribution over CT attributions at the lateral cervical site. These findings agree with our prior observations on unimodal LNM prediction performance, highlighting a more prominent role for US at the central site, whereas both US and CT show comparable importance at the lateral cervical site. In addition, this underscores that the DGFNet model can dynamically adjust the weights of the two modalities based on nodule characteristics, effectively leveraging the strengths of both modalities.

### DGFNet enhances model attention on the nodular region in US and CT images

To further investigate how our DGFNet model improves the LNM prediction performance, we generated saliency maps for both US and CT images in the multimodal network and compared them with their unimodal counterparts. The results clearly show that, for both US and CT images, the DGFNet model significantly increases the attention towards the region of interest compared to the unimodal models. Specifically, within US images, the multimodal model focuses more intensely on the nodules’ peripheral and inner hypoechoic region (Fig. 4a), whereas in CT images, it narrows its focus to the nodules and their immediate surrounding tissues (Fig. 4b), all of which represent crucial regions providing key information for LNM prediction. This directly proves the superiority of DGFNet in grasping meaningful medical information over unimodal methods.

**Figure 4:**
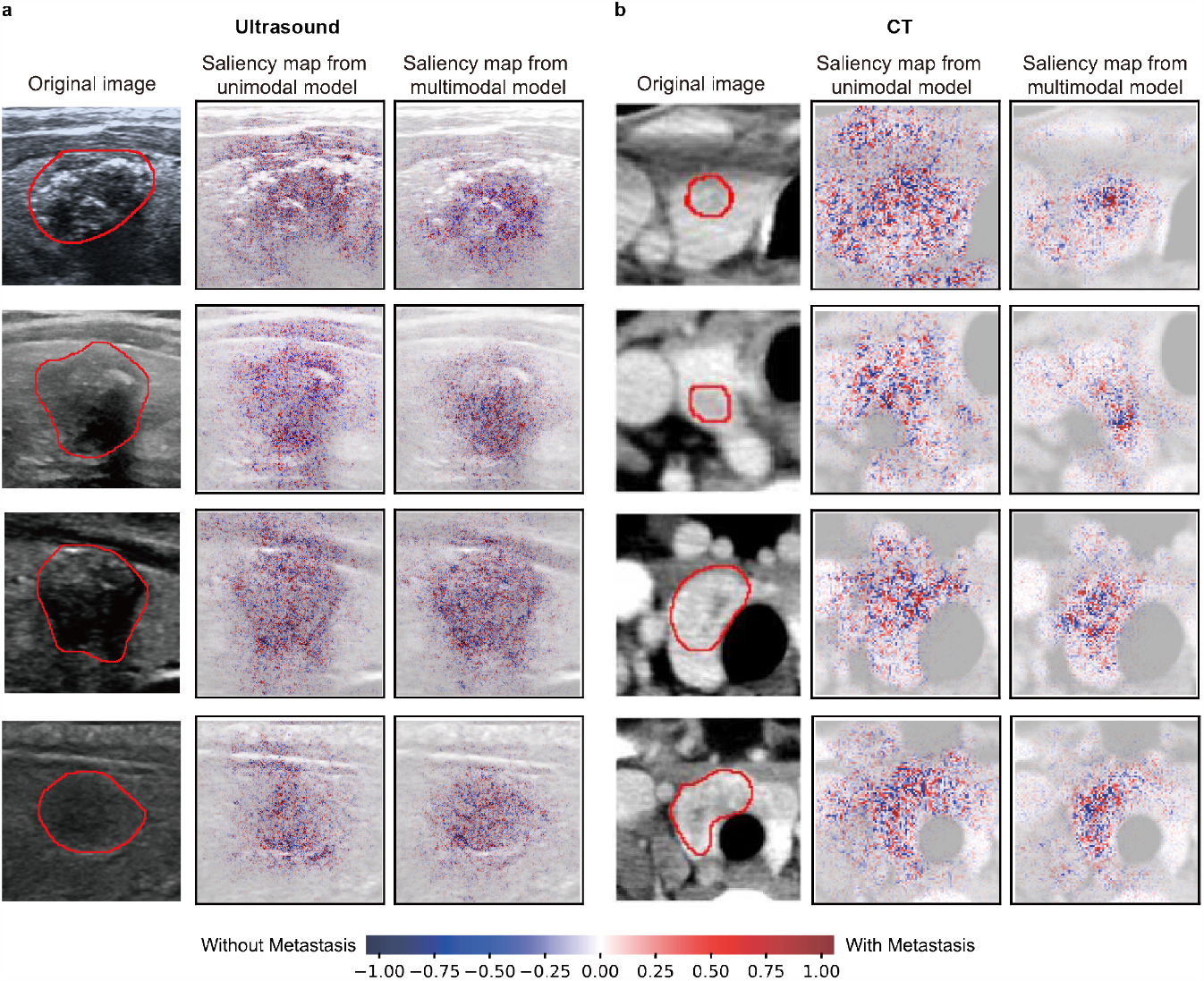
Examples of saliency map visualization results at central site. In these instances, both the US and CT unimodal models initially generated inaccurate predictions, whereas the multimodal models effectively rectified these to provide accurate predictions. The red curve delineates the nodule’s boundary in the original US and CT images. The color red signifies an elevated likelihood of LNM development, whereas the color blue signifies the contrary.

### Identify patients who can best benefit from multimodal integration

The multimodal approach can effectively improve the LNM prediction, however, it is often unfeasible to examine all patients by both modalities in real clinical settings. Hence, to make our DGFNet more applicable and useful for clinicians, we further seek to identify patients who can best benefit from the multimodal approach. Given that the US examination is cheaper and more commonly used, we analyzed by identifying cases for whom adding CT as a supplementary modality would be advantageous. We evaluated four well-established sonographic characteristics of the thyroid nodule during US diagnosis including maximum diameter, margin characteristics, aspect ratio, plus location in the thyroid for central cite nodules, and categorized the nodules based on the measurements. We then compared the prediction performance in each category between the DGFNet and the unimodal model of US and CT respectively. (Results on more characteristics are illustrated in Supplementary Material)

The analysis shows that the DGFNet model achieves particularly high performance in specific circumstances. This includes nodules with maximal diameters between 20mm and 36mm, as well as more extreme cases less than 12mm or larger than 60mm(Fig. 5a). Additionally, DGFNet excels in cases exhibiting non-smooth borders (Fig. 5b), aspect ratios surpassing 1 (Fig. 5c), and nodules situated within the thyroid isthmus (Fig. 5d). Similar findings are observed in the analysis conducted at the lateral cervical site (Supplementary Material). Therefore, the DGFNet model is potentially particularly beneficial and practical for patients with the above nodule characteristics.

**Figure 5:**
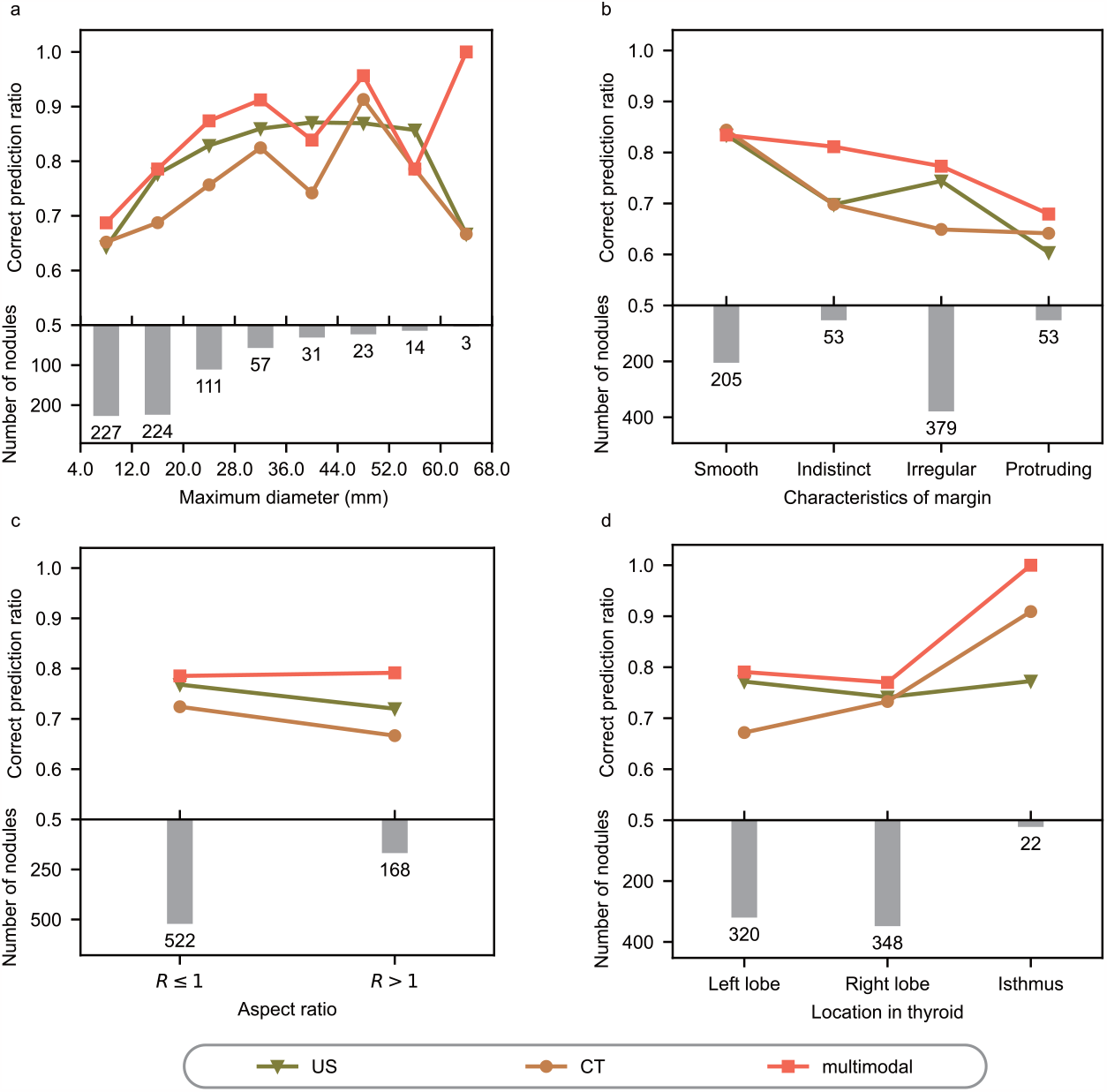
Distribution of nodules with varied attributes and associated correct predictions ratio in central Site. Attributes encompass nodal maximum diameter (considering the larger of the maximum diameters from transverse and longitudinal US views) in US image(a), characteristics of margin (b), aspect ratio (calculated as the height divided by the width in transverse views) of nodules(c), and location in thyroid (d).

## Discussion

Patients often undergo multiple types of examinations in the diagnostic process, and the effective integration of multimodal information can greatly improve diagnosis accuracy. Recent advancements in artificial intelligence techniques have facilitated the progress of deep-learning-based multimodal integration methods, which have emerged as a trend in cancer diagnosis in recent years. In this study, we pioneered the development of a multimodal deep learning approach that effectively integrates US and CT modalities to successfully enhance the accuracy of LNM prediction and further demonstrate its generalizability in an external dataset. Moreover, by conducting a series of comprehensive interpretability analyses, we quantified the modality-specific contribution across nodules in various situations, and investigated the attention heatmap of US and CT images within the model, which not only shed light on the reasons for the improved performance of the multimodal model, but also improve the model’s applicability in clinical settings, and opens a new avenue for mitigating the problem of overtreating thyroid cancer.

The effective integration of multimodal data often relies on a deep understanding of the domain knowledge involved with specific medical tasks. In our study, a close collaboration between AI scientists and clinicians allowed us to leverage our collective expertise in deep learning models, thyroid cancer, US and CT images. This enabled us to strategically employ multi-task learning techniques, facilitating the identification of critical regions and extraction of essential LNM-related features from both US and CT images. Moreover, we introduced a novel diverse granularity fusion network (DGFNet) that learns the attention from three different levels, which excels in not only effectively integrating shared and specific features from multimodal data but also dynamically adjusting the weights of each modality’s data for different nodules. This approach demonstrates the potential to optimize the utility of both US and CT images and aggregate information from similar nodules, thereby enhancing the model’s overall performance and robustness.

Besides the excellent performance of our developed DGFNet model, our study has yielded valuable clinical insights through the multimodal approach. First, it shows that unimodal methods based on US appear to be more sensitive but less specific, while CT-based unimodal methods are the other way around. Second, it shows that the US modality generally plays a more significant role than CT at the central site, whereas there is no obvious difference between US and CT at the lateral cervical site. Furthermore, by quantifying the performance of the unimodal and multimodal models for nodules within different diagnosis characteristics categories, we could pinpoint patients with certain nodule characteristics who can potentially best benefit from the multimodal approach. These analyses offer valuable insights for accurately identifying the appropriate patient population for multimodal diagnostic approaches in clinical practice and guiding patients in selecting the most suitable examination method.

In conclusion, through a close collaboration between AI scientists and clinicians, this study successfully develops a multimodal approach aimed at improving the LNM prediction for thyroid patients. It paves the way for addressing the issue of overtreatment in thyroid cancer and provides new insights in the integration of multimodal data for precise diagnosis, representing an excellent scientific research example originating from clinical practice and directly addressing clinical necessities.

## Methods

### Patient Cohort

There are two cohorts included in this study. The main cohort was obtained from Zhejiang Cancer Hospital in Zhejiang Province, China, consisting of 1360 patients. After the data screening process, a total of 1138 patients with 1285 nodules were retained for analysis. The main cohort was utilized for the model establishment and internal performance evaluation. The second cohort, referred to as the external cohort, was sourced from Shaoxing People’s Hospital in Zhejiang Province, China. Initially, this cohort included 126 patients, and after the data screening process, 60 patients with complete data were included for evaluation of model generalization (The patient enrollment process is illustrated in Supplementary Material). Ethical approval for the study was obtained from the ethics committees of both hospitals and verbal informed consent was obtained from all participating patients.

The inclusion criteria for this study encompassed the following: (1) patients with thyroid nodules, (2) patients who underwent cervical US and CT examinations, and (3) patients with confirmed pathological status of cervical LNM. Exclusion criteria consisted of the following: (1) missing US or CT data, (2) the presence of measurement lines in the US images, and (3) patients with multiple malignant thyroid nodules and metastatic cervical lymph nodes. As all the thyroid nodules had the potential to metastasize, it was impossible to determine which specific nodule had metastasized to the cervical lymph nodes. After the data screening process, the number of nodules with and without metastasis in the central and lateral cervical sites for both cohorts is presented in the Supplementary Material.

Multiple US images, including transverse and longitudinal sections, as well as multiple CT images from different slices, were available for most nodules in the dataset. During each epoch of the training process, one random US image and one random CT image were paired together to form an image pair. During the evaluation process, the US and CT images with the largest nodal area were selected from the multiple available images to form an image pair for analysis.

### Data pre-processing

#### Region of Interest Extraction

The methods used for extracting the region of interest in both US and CT images are similar and described as follows: 1) We first performed a dilation operation on the mask of thyroid nodules annotated by clinicians, using a 3×3 dilation kernel. The iteration steps were set to 40 and 25 for US and CT images, respectively. 2) We determined the horizontal bounding rectangle of the dilated region, with the height, width, and center coordinates of the rectangle denoted as *h, w*, and (*x*_center_, *y*_center_), respectively. 3) Using (*x*_center_, *y*_center_) as the center and the larger value of *h* and *w* as the side length, we obtained the external square of the thyroid nodule. 4) The original US and CT images were then cropped to reserve the region within this square. If the square area exceeds the image boundary, the images are padded with zeros to fill the exceeding part.

#### Image Augmentation

The cropped US and CT images were resized to 288×288 pixels and 96×96 pixels, respectively. To enhance the diversity of the data, we applied additional data augmentation techniques to both modalities. These techniques included rotation, horizontal flip, cropping and scaling, brightness-contrast transformation, and elastic transformation. For rotation, the angle of rotation ranged from -15° to 15°. Random cropping occurred with the cropped area set to be between 90% and 100% of the original size. The probability of applying these transformations was set to 0.5, ensuring a balanced augmentation effect.

### Convolutional neural network architecture

The architecture of the proposed model is depicted in Fig. 2. The model is composed of three distinct branches: the US branch, the CT branch, and the Multimodal branch. Both the US and CT branches share an identical structure, each comprising an encoder and two decoders. The encoder adopts a pre-trained ResNet[34] architecture, with ResNet34 and ResNet18 selected for the central and lateral cervical sites, respectively. Regarding the US branch, the decoders are trained to delineate the mask and boundary of thyroid nodules, directing the model’s attention towards the internal and marginal regions of the nodules, correspondingly. Conversely, the CT branch’s decoders focus on segmenting the mask of thyroid nodules and the boundary of surrounding tissue, facilitating the model in comprehensively capturing information about both the thyroid nodule and its adjacent surroundings.

All the aforementioned decoders share the same structure. Each decoder is constructed from 5 upsample blocks, with every block encompassing 2 layers. In the initial layer of each block, the input feature is upsampled using bilinear interpolation. Subsequently, the second layer comprises a convolutional block, incorporating a convolutional layer featuring a kernel size of 3×3, followed by batch normalization, relu activation, and a dropout layer. Notably, to enhance segmentation performance, short connections interconnect the encoder and decoder components.

The US and CT encoders produce 512-dimensional vectors through global average pooling. In unimodal models, these vectors directly enter the classifier for LNM prediction. In the multimodal model, the unimodal vectors integrate within the multimodal branch and then proceed to the classifier with the same structure—a two-layer fully connected neural network with 512 input nodes and a single output node.

### Diverse granularity fusion module

The diverse granularity fusion module comprises three branches, as depicted in the supplementary material. All branches are constructed using the attention mechanism.

In the dimensional correlation branch, the US and CT features undergo a preliminary transformation as outlined below:

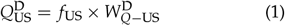

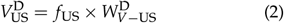

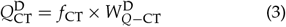

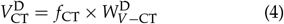

Here, *f*_US_ and *f*_CT_ are the unimodal features of US and CT, respectively, and 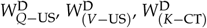 and 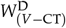 are trainable parameters. The product of 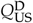 and 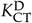, followed by the application of the softmax function, results in the attention matrix *A*^D^, which captures the interplay between various feature dimensions of the US and CT modalities:

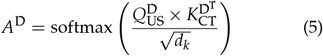

Here, *d*_*k*_ is the dimension of 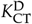 . The derived attention matrix is then utilized for the enhanced multimodal features:

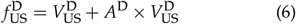

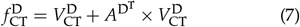

Ultimately, the enriched features are amalgamated through concatenation along the dimension axis, yielding the fused features:

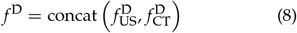

In the modal weights branch, the US and CT features are first concatenated along the modal axis:

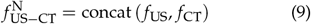

Then the *Q*^M^, *K*^M^, and *V*^M^ are generated respectively:

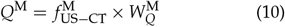

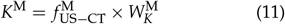

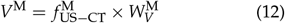

The 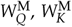, and 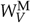 are trainable parameters. Through the multiplication of *Q*^M^ and *K*^M^, an attention matrix emerges, encapsulating the priority of the two modalities within separate nodes.

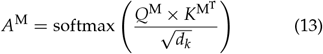

Subsequently, this attention matrix is employed to adjust the relative significance of the two modalities:

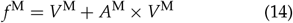

In the nodal correlation branch, US and CT features are first merged along the dimensional axis:

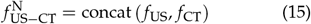

Then *Q*^N^, *K*^N^, and *V*^N^ are obtained respectively:

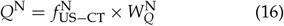

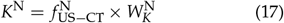

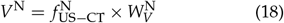

The 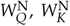, and 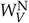 are trainable parameters. The attention matrix is obtained and employed to delineate the interrelation between distinct nodules:

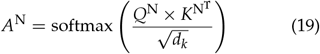

Refined features considering the similarity of different nodules emerge:

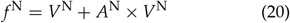

The features from the three branches undergo element-wise multiplication, resulting in the ultimate fused features:

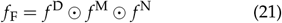

### Nodule boundary extraction in US images

Firstly, the nodule boundary width (*d*) was determined as a multiple ( *f* ) of the square region of interest’s length. For our study, *f* was set to 0.08. Secondly, the annotated thyroid nodule mask underwent dilation and erosion operations to yield *R*_dilation_ and *R*_erosion_, respectively, with a kernel size of 3×3 and iterations of 0.5*d*. Finally, the nodule’s boundary was obtained as the difference between *R*_dilation_ and *R*_erosion_ (*R*_dilation_ − *R*_erosion_).

### Boundaries of surrounding tissue extraction in CT images

Firstly, bilateral filtering[36] was applied to preserve the edges while reducing noise. The diameter of the pixel field was set to 7, and the sigma values for both the color space and coordinate space were set to 100. Secondly, the Canny algorithm[37] was employed to further extract the boundaries of the surrounding tissues. The lower and upper threshold values were set to -100 and 200, respectively.

### Training Configuration

The base learning rate in our study was set to 1*×* 10^−4^, and we employed a cosine learning rate schedule during the training process. The batch size was set to 30, and we utilized the Adam optimizer to optimize our model. A weight decay of 1 *×* 10^−5^ was applied to mitigate overfitting. In this study, a multi-task strategy was employed to address different tasks. For the classification task, specifically the prediction of the LNM status, we utilized a binary cross-entropy loss function. As for the segmentation tasks, a combination of binary cross-entropy loss and Intersection over Union (IOU) loss functions was utilized. The model was initially trained for the segmentation tasks for the first 100 epochs, and then the classification task was added and trained for the remaining 200 epochs.

### Interpretability Analysis Methods

We employed the integrated gradients[35] method to enhance the interpretability of our model. Integrated gradients is a feature attribution technique that calculates the integral of gradients along the path from a chosen baseline to the input, resulting in an attribution value for each input feature. In our study, the baseline is manually specified, and we select a baseline where the predicted probability of our trained model is close to 0.5, indicating equal probabilities for both LNM presence and absence. To determine the contributions of US and CT images, we sum the attributions of each pixel in the respective images. By visualizing the attribution of each pixel, we generate saliency maps for US and CT images.

### Statistical Analysis

We assessed the performance of our model using several evaluation metrics, including accuracy, area under the curve (AUC), specificity, sensitivity (also known as recall), precision, and F1-score. To analyze the model’s performance across different thresholds, we constructed receiver operating characteristic (ROC) curves, plotting sensitivity against specificity.

### Hardware and Software

The computational resources utilized include an Intel 10900K CPU with a clock speed of 3.7GHz and 20 threads. The graphics card employed is a GEFORCE RTX 3090, equipped with 10752 CUDA cores and 24GB of graphics memory. The programming language used for implementation is Python 3.9.7, and the deep learning framework employed is PyTorch 1.10.0.

## Data availability

Though this study was carried out with participant consent, the dataset remains restricted in public access. For research inquiries, de-identified data can be obtained from the corresponding author upon reasonable request.

## Code availability

The code for model development and interpretability analysis is accessible at https://github.com/li10107/DGFNet.

## Acknowledgments

This work was supported by the National Natural Science Foundation of China (No. 82071946), the Natural Science Foundation of Zhejiang Province (No. LZY21F030001), the Pioneer and Leading Goose R&D Program of Zhejiang (No. 2023C04039), the National Key Research and Development Program of China (2022YFF0608403), Youth Research Fund Project of Shaoxing People’s Hospital (Grant Number 2022YB07), and the fund of Zhejiang Province Medical and Health Science and Technology Project (No. 2023KY581). We thank Y.G. for providing us external validation set.

## Author contributions

M.H., C.S., X.L., and D.X. conceived and planned the study. C.S., G.L., and X.L. designed the research framework. J.Y., C.P., S.Z., and J.Y. collected the raw US and CT images, patients’ clinical information, and image annotation. G.L., Y.H., and X.F. performed the data preprocessing and conducted the performance analysis. G.L. designed the multimodal fusion method and carried out model interpretation analysis. G.L. and C.S. wrote the manuscript. All authors commented on the manuscript.

