## Supplementary Material for "Multimodal Diverse Granularity Fusion Network based on US and CT Images for Lymph Node Metastasis Prediction of Thyroid Carcinoma"

**Table 1:** *Number of positive and negative nodules in central and lateral cervical sites. Positive and negative mean the presence and absence of LNM respectively.*

| | Central site | | Lateral cervical site | |
|---|---|---|---|---|
| | Negative | Positive | Negative | Positive |
| Main cohort | 808 | 477 | 1084 | 201 |
| External cohort | 38 | 22 | | |

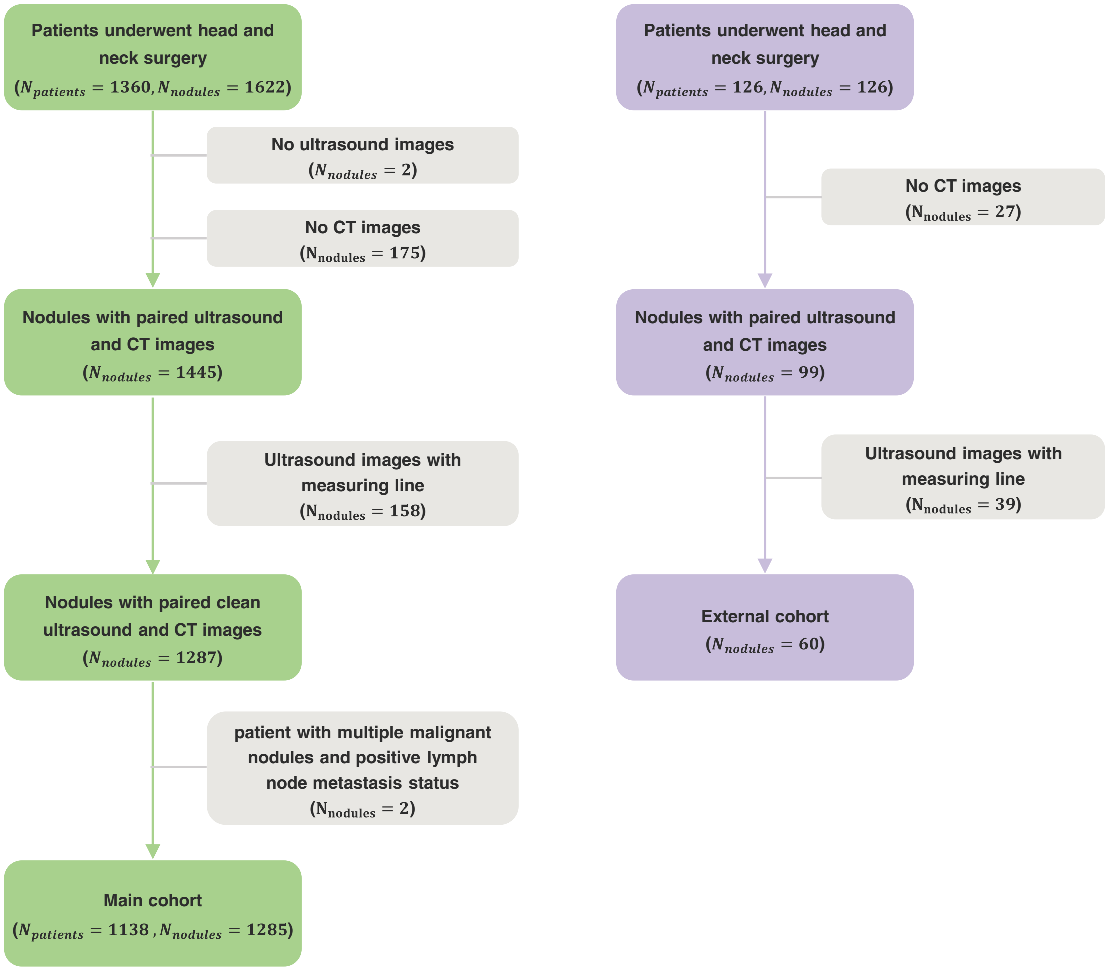


**Figure 1:** *Patient enrollment process for main and external cohorts. The diagram illustrates the patient enrollment procedures for both the main cohort (left side) and the external cohort (right side).*

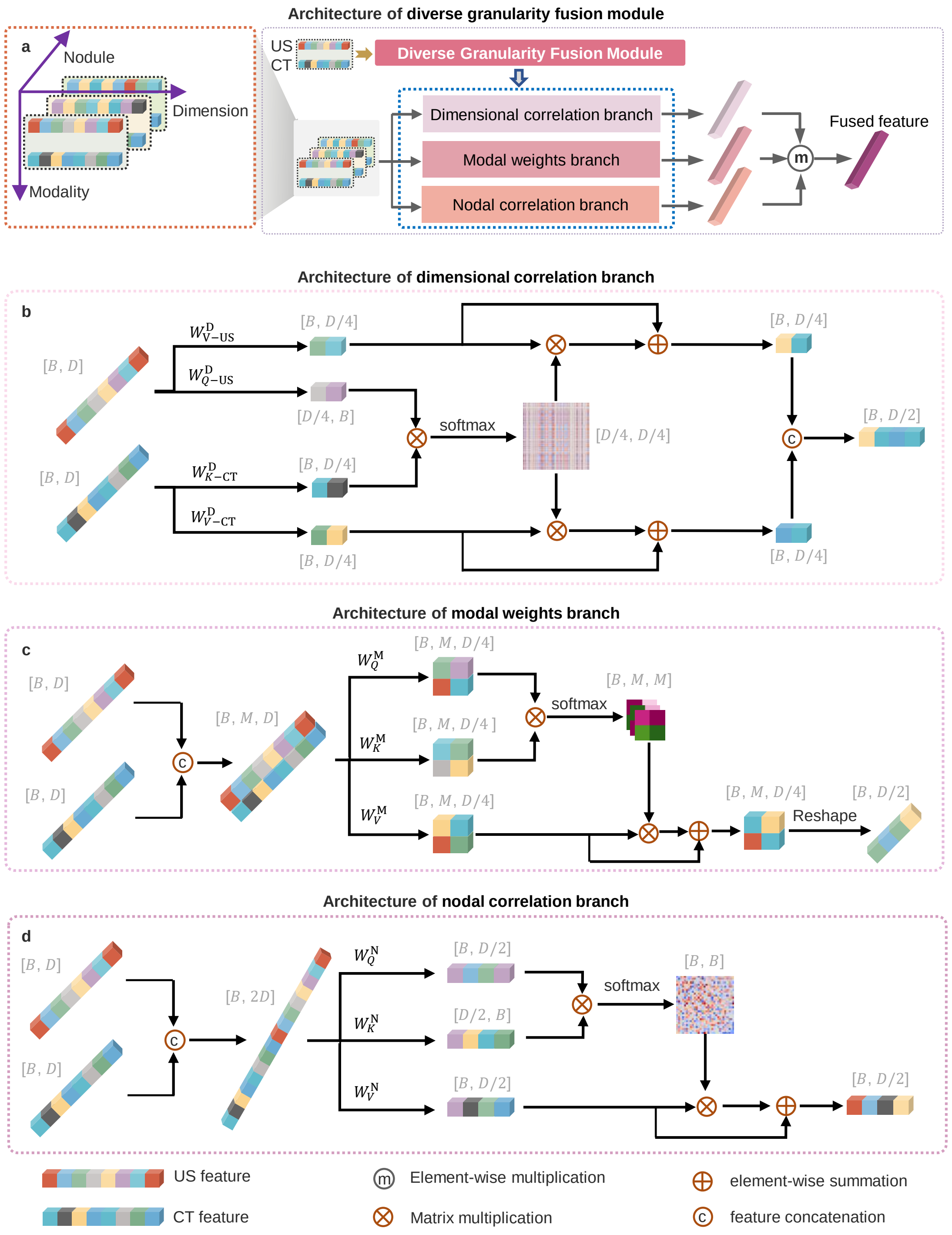


**Figure 2:** *Structure of diverse granularity fusion module (a), dimensional correlation branch (b), modal weights branch (c), and nodal correlation branch (d). B, D, and M represent batch size, number of dimensions, and number of modalities respectively.*

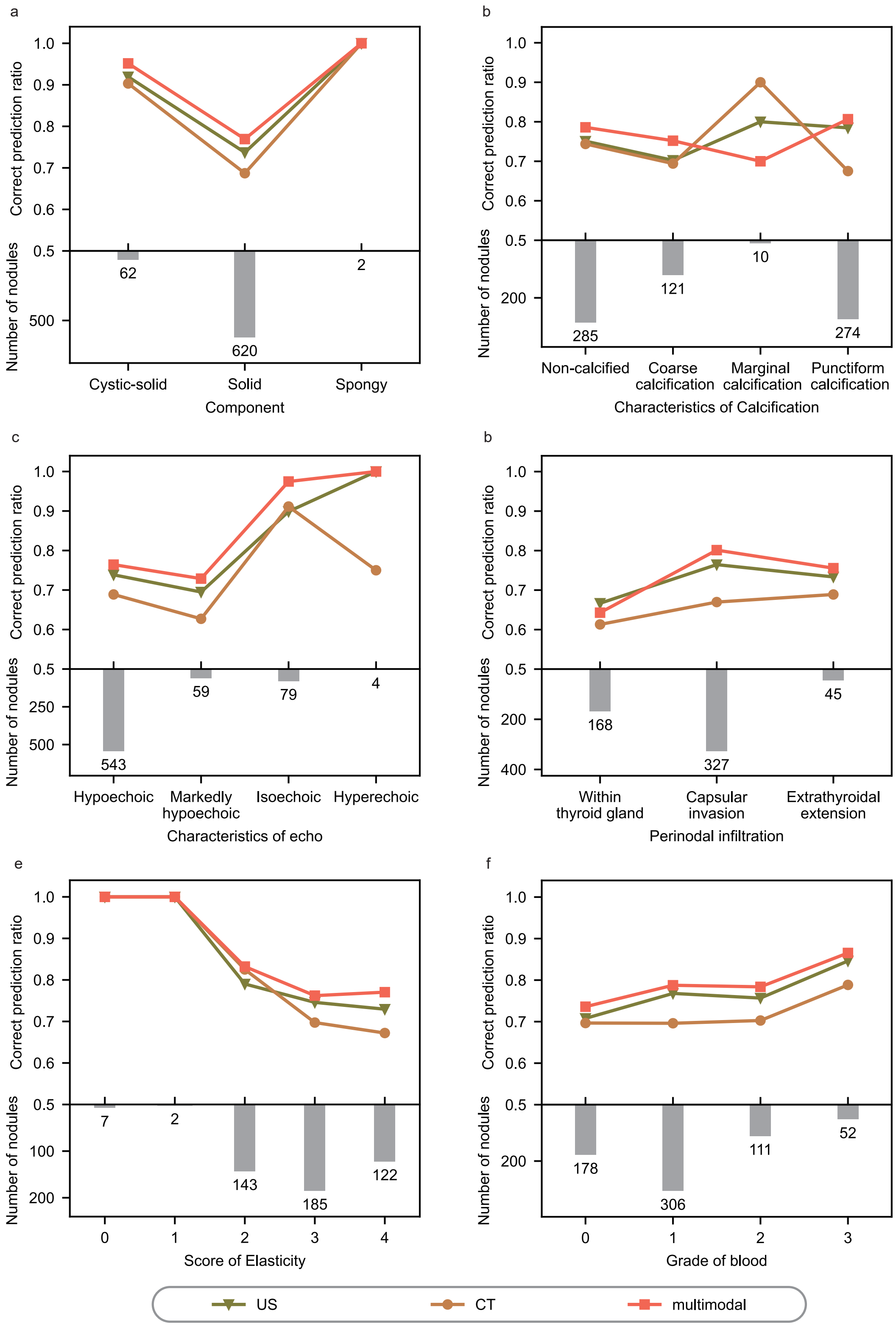


**Figure 3:** *Distribution of nodules with varied attributes and associated correct predictions ratio in central site. Attributes encompass components of nodules(a), characteristics of calcifications (b), characteristics of echo (c), perinodal infiltration (d), score of elasticity[38] (e), and grade of blood[39].*

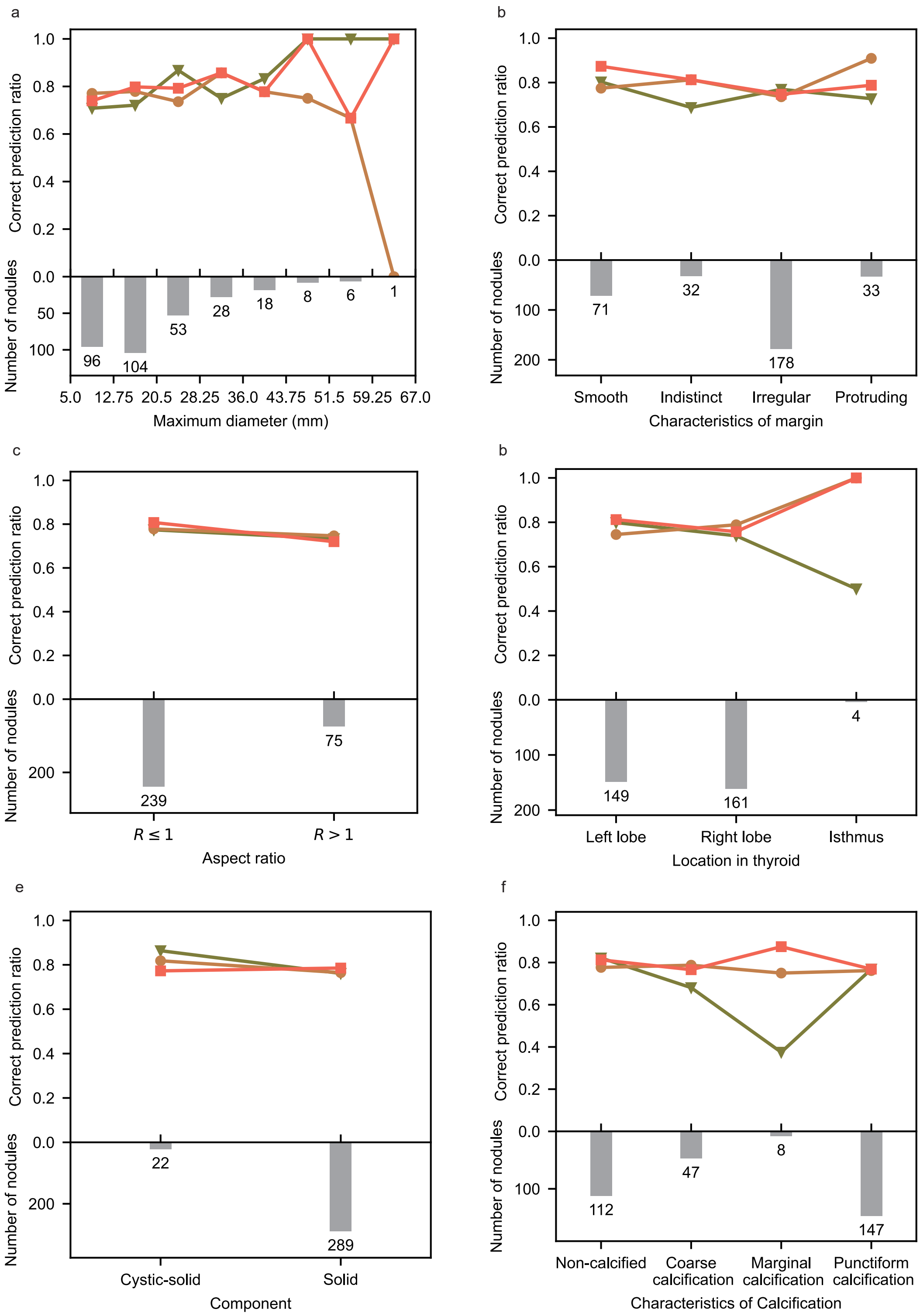
a
Correct prediction ratio
Number of nodules
96
104
53
28
18
8
6
1
5.0 12.75 20.5 28.25 36.0 43.75 51.5 59.25 67.0
Maximum diameter (mm)
b
71
32
178
33
Smooth
Indistinct
Irregular
Protruding
Characteristics of margin
c
239
75
$R \leq 1$
$R > 1$
Aspect ratio
b
149
161
4
Left lobe
Right lobe
Isthmus
Location in thyroid
e
22
289
Cystic-solid
Solid
Component
f
112
47
8
147
Non-calcified
Coarse calcification
Marginal calcification
Punctiform calcification
Characteristics of Calcification

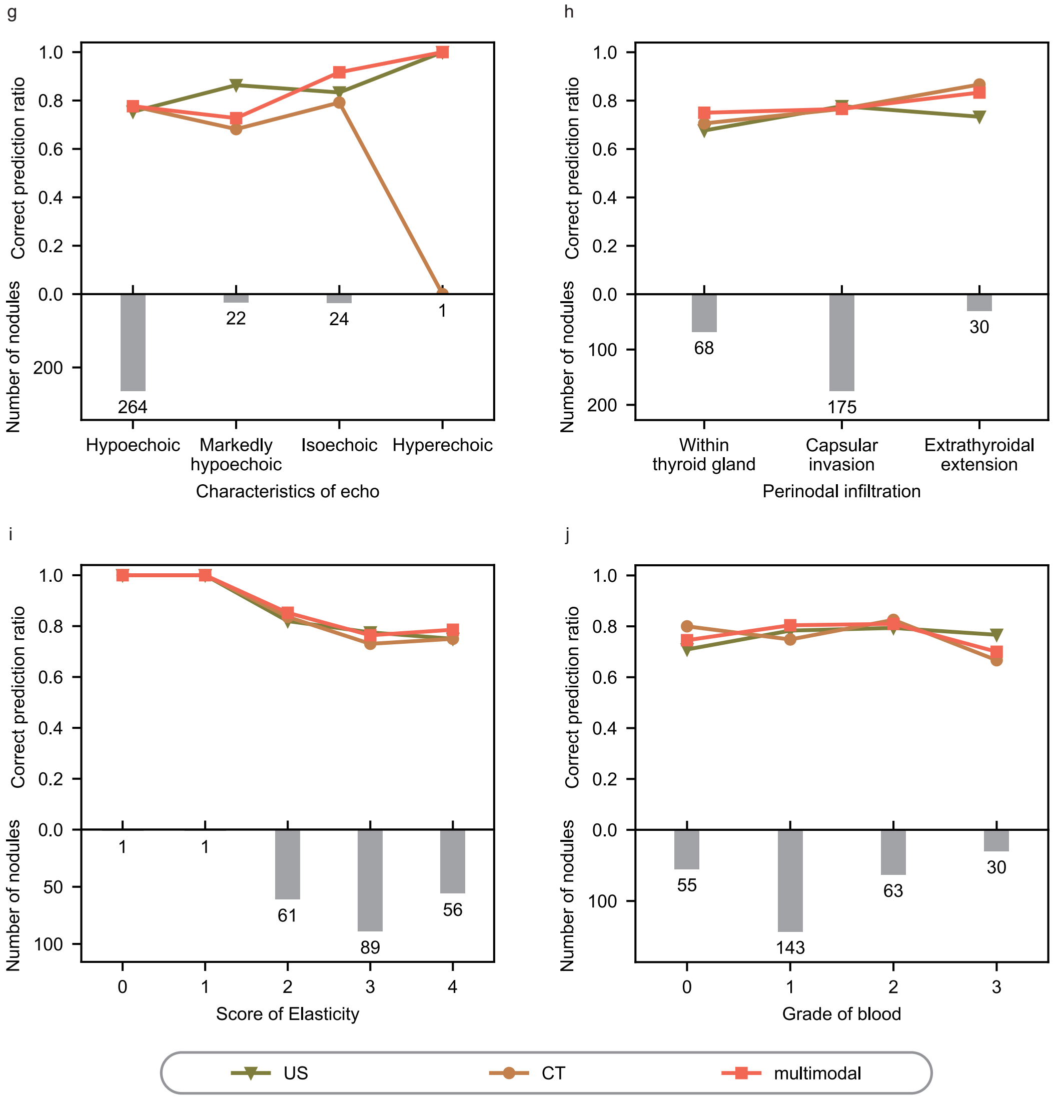


**Figure 4:** *Distribution of nodules with varied attributes and associated correct predictions ratio in lateral cervical site. Attributes encompass nodal maximum diameter in US image(a), characteristics of margin (b), aspect ratio of nodules(c), location in thyroid (d), components of nodules(e), characteristics of calcifications (f), characteristics of echo (g), perinodal infiltration (h), scores of elasticity (i), and grade of blood (j).*
